# Automating the Clinical Interpretation of *LDLR, APOB*, and *PCSK9* Variants: A Web-Based Platform for FH Diagnosis

**DOI:** 10.64898/2026.01.10.26343831

**Authors:** Helene Gellert-Kristensen, Tim Møller Eyrich, Stefan Stender

## Abstract

**Background:** Clinical interpretation of *LDLR, APOB*, and *PCSK9* variants is essential for diagnosing familial hypercholesterolemia (FH), yet manual evidence curation is labor-intensive and prone to variability.

**Methods:** We developed a web-based platform (**https://fh-interpret.shinyapps.io/beta/**) to automate the synthesis of multi-dimensional evidence. The tool integrates population frequencies (gnomAD), phenotype associations (UK Biobank), AI-based pathogenicity scores (AlphaMissense), high-throughput functional data from saturation mutagenesis, and established clinical classifications (ClinVar, HGMD).

**Results:** The application streamlines the diagnostic workflow by automatically synthesizing: (1) population frequencies from gnomAD to assess variant rarity; (2) statin-adjusted LDL-cholesterol association analyses in ∼470,000 UK Biobank participants; (3) AlphaMissense pathogenicity predictions; (4) functional scores for ∼17,000 LDLR variants quantifying impact on LDL uptake and LDLR cell-surface abundance; (5) existing clinical evidence from ClinVar and HGMD; and (6) identification of pathogenic variants at identical amino acid positions (ACMG PM5). The tool generates narrative evidence summaries to facilitate standardized ACMG/AMP classification.

**Conclusions:** By providing real-time access to massive-scale genomic, phenotypic, and functional datasets within a single platform, this tool streamlines the FH diagnostic workflow and supports more consistent and efficient clinical variant interpretation.

**Graphical Abstract:** 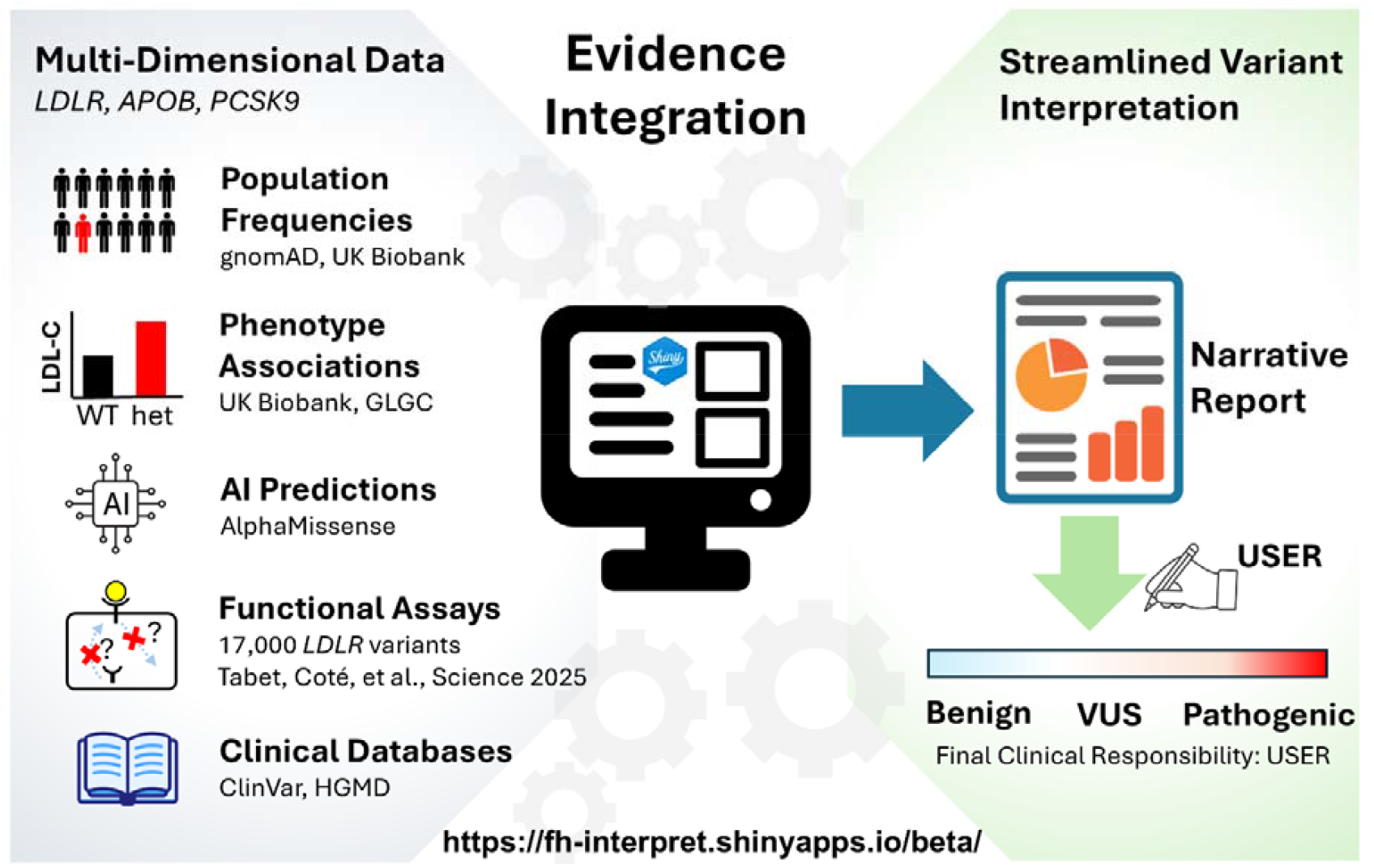

## Background

Familial hypercholesterolemia (FH) is a common autosomal dominant disorder characterized by lifelong high blood concenstration of low-density lipoprotein cholesterol (LDL-C)[1–3]. If left untreated, FH leads to a significantly increased risk of premature atherosclerotic cardiovascular disease. With a global prevalence estimated at 1 in 250 individuals, FH remains a major public health challenge and is profoundly underdiagnosed[1].

Genetic testing is a cornerstone of FH diagnosis, typically focusing on *LDLR, APOB*, and *PCSK9*. While high-throughput sequencing has improved mutation identification, it has created a “bottleneck” in variant interpretation. The current ACMG/AMP standards require integrating population frequency, computational predictions, functional assays, and genotype-phenotype correlations[4].

Currently, this data is fragmented across resources like gnomAD, ClinVar, and specialized literature. Manually gathering and evaluating this evidence is labor-intensive and susceptible to human error. To address these limitations, we developed an integrated, web-based platform to automate the FH evidence-gathering workflow, providing a “one-stop” resource to reduce interpretation turnaround time and improve diagnostic yield.

### Application Development

We developed a web-based application using R (version 4.3.2) and Shiny for R (version 1.11.1)[5,6]. The platform aggregates multi-dimensional evidence to generate standardized draft interpretation summaries. The application is designed to streamline evidence curation; however, all generated text is intended as a preliminary draft. Final clinical classifications remain the sole responsibility of the user. Integrated data sources include:

1. **Population Allele Frequencies:** Data is retrieved from gnomAD v4.1.0 (∼1.6 million alleles)[7].
2. **UK Biobank (UKB) Phenotype Associations:** Associations with LDL-C were assessed in ∼470,000 participants[8]. Participants on statins had their measured LDL-C multiplied by 1.42 to correct for the statin-induced reduction in LDL-C of an average of 30%. Statistical associations were evaluated using linear regression adjusted for age, sex, and the first ten principal components of ancestry. Specific counts for variants observed in <5 individuals are masked per UKB guidelines.
3. **GWAS Meta-Analysis (GLGC):** Data from the 2017 GLGC exome chip GWAS (∼300,000 individuals) provide independent per-allele effects on LDL-C[9].
4. ***In Silico* Pathogenicity (AlphaMissense)[10]:** Predictions were sourced for *LDLR* and *PCSK9*, utilizing recommended thresholds: <0.34 (benign), 0.34–0.56 (uncertain), and >0.56 (likely pathogenic).
5. **Clinical Databases:**
  - HGMD[11]: Includes missense, nonsense, and splice variants associated with FH (Class ‘DM’).
  - ClinVar[12]: Extracted FH-associated variants for *LDLR* and *PCSK9*.
6. **Amino Acid Position (ACMG PM5):** The app identifies other missense variants at the same position classified as pathogenic in HGMD or ClinVar.
7. **High-Throughput Functional Assays:** Evidence is derived from a 2025 deep mutational scanning study of ∼17,000 *LDLR* variants[13]. Scores <0.5 of wild-type function are defined as impaired.

### Application Functionality and Case Studies

The application interface is organized into four parts that facilitate real-time data exploration (see screenshot below). The top left panel allows for the selection of the gene of interest. A table at the bottom includes all variants within the selected gene. Specific variants can be identified via a global search field (e.g., by nucleotide or amino acid). Furthermore, columns within the table can be ordered to facilitate data-driven discovery—for example, by sorting the p-value column to prioritize variants with the most statistically significant phenotypic effects.

The central graph displays mean statin-adjusted LDL-C concentrations (95% CI) stratified by genotype within the UK Biobank for the selected variant. A narrative interpretation that integrates all available evidence for the selected variant is shown above the graph. Selecting a variant in the bottom table automatically refreshes the interpretation and central graph.

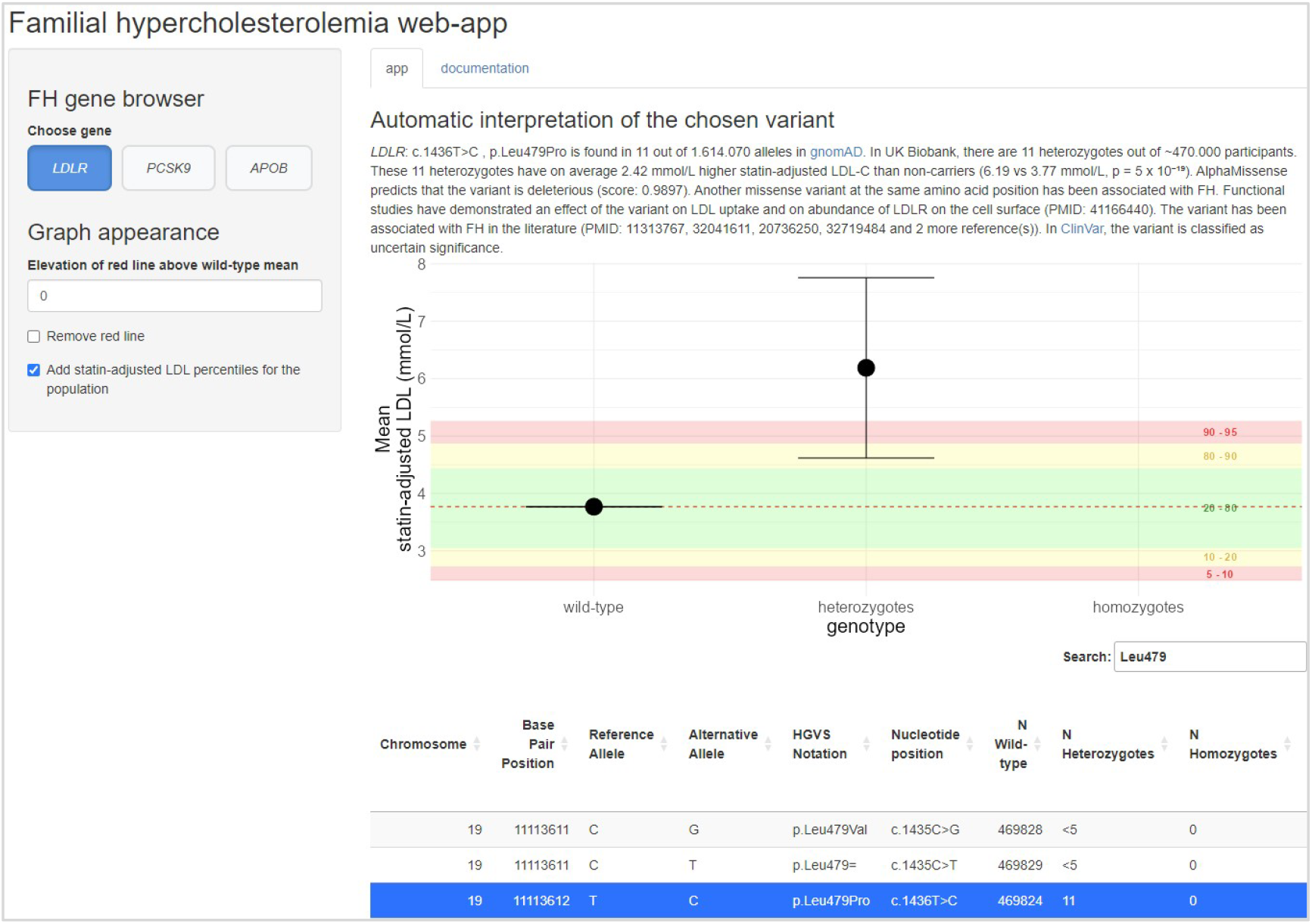

#### Case Study 1: Reclassifying LDLR p.Leu479Pro

The *LDLR* p.Leu479Pro variant was recently classified as a VUS by the ClinGen FH Variant Curation Expert Panel (April 2025)[14]. However, our application aggregates multidimensional evidence suggesting pathogenicity:

- **UKB phenotype:** 11 heterozygotes exhibited a mean statin-adjusted LDL-C 2.42 mmol/L higher than non-carriers (6.19 vs 3.77 mmol/L; p = 5 × 10^-19^), providing strong evidence of a large clinical effect.
- ***In Silico*:** AlphaMissense score of 0.99 (deleterious).
- **Functional data:** High-throughput functional assays demonstrated impaired LDL uptake and LDLR cell-surface abundance.

The robust UKB association data, integrated alongside functional and *in silico* evidence, supports a reclassification of this variant to pathogenic.

#### Case Study 2: Evidence for Benign Classification of LDLR p.Glu380Lys

*LDLR*: p.Glu380Lys is currently a VUS in ClinVar. Despite being located at a position where another variant is pathogenic, our tool suggests it is benign:

- **UKB:** 29 heterozygotes have a mean LDL-C nearly identical to non-carriers (3.73 vs 3.77 mmol/L; p = 0.86).
- ***In Silico*:** AlphaMissense score of 0.07 (likely benign).
- **Functional:** No deleterious effect on LDL uptake or LDLR cell-surface abundance.

The lack of LDL-C elevation in 29 heterozygotes provides strong evidence against a large effect size, preventing “guilt by association” misinterpretation.

## Discussion

The interpretation of variants in *LDLR, APOB*, and *PCSK9* is the primary rate-limiting step in the molecular diagnosis of FH. By consolidating population-scale biobank data, AI-driven pathogenicity scores, and high-throughput functional assays into a single interface, our application addresses the inherent inefficiencies of manual evidence curation.

Despite its utility, the application has limitations. The UK Biobank population is predominantly of European ancestry, which may limit the generalizability of the phenotypic evidence for variants found more frequently in other global populations. Future iterations of the tool should aim to incorporate data from more diverse cohorts, such as the All of Us Research Program[15]. Furthermore, while the tool automates the retrieval of evidence, it is designed to support — not replace — clinical judgment. The final classification must still be made by a qualified professional who can weigh the automated summary evidence against the specific clinical context of the patient.

In conclusion, our web-based application provides a “one-stop” digital ecosystem for FH variant interpretation. It empowers clinicians and academics to interpret genetic variation with greater speed, consistency, and evidence-based confidence. As genomic medicine continues to scale, such tools will be essential for realizing the full potential of precision cardiovascular care.

## Data Availability

The application is open-access at https://fh-interpret.shinyapps.io/beta/. Source code and data is available on GitHub at https://github.com/tme34/FH-interpret under the MIT License.

## Conflicts of interest

All authors declare no competing interests.

## Author contributions

*Helene Gellert-Kristensen:* Conceptualization, Software (UI design and R-code development), Visualization, Writing - Review & Editing. *Tim Møller Eyrich:* Data curation, Formal analysis (UK Biobank), Software (algorithm logic and R-code development), Writing - Review & Editing. *Stefan Stender:* Conceptualization, Funding acquisition, Supervision, Writing - Original Draft.

## Funding statement

This work was supported by Independent Research Fund Denmark (Sapere Aude Research Leader 9060-00012B) and the Novo Nordisk Foundation (Borregaard Clinical Ascending Investigator NNF22OC0075038). The funders played no role in study design, data collection, analysis and interpretation of data, or the writing of this manuscript.

## Data availability

The application is open-access at **https://fh-interpret.shinyapps.io/beta/**. Source code and data is available on GitHub at **https://github.com/tme34/FH-interpret** under the MIT License.

## Acknowledgements

This research used the UK Biobank Resource (Application 104807). The authors acknowledge the use of Google Gemini for linguistic refinement and readability improvements; final content and scientific integrity remain the responsibility of the authors.

## Ethics

The UKB study was approved by the North West Multi-centre Research Ethics Committee, and all participants provided written informed consent to participate in the UKB. All other data included in the application are derived from previously published, publicly available datasets. These studies received ethical approval from their respective local or national institutional review boards.

## Notes

### Competing Interest Statement

The authors have declared no competing interest.

### Author Declarations

This research used the UK Biobank Resource (Application 104807). The UKB study was approved by the North West Multi-centre Research Ethics Committee, and all participants provided written informed consent to participate in the UKB. All other data included in the application are derived from previously published, publicly available datasets. These studies received ethical approval from their respective local or national institutional review boards.

